# What are the barriers and facilitators to women with menopausal symptoms participating in physiotherapy intervention?

**DOI:** 10.1101/2025.07.15.25331542

**Authors:** Julia Gross, Gareth Stephens, Andrew Soundy

**Affiliations:** Royal Orthopaedic Hospital, Bristol Road South, Northfield, Birmingham B31 2AP UK; University of Birmingham, School of Sport, Exercise and Rehabilitation Sciences, Y14, Edgbaston, Birmingham B15 2TT UK

**Author notes:** <u>Corresponding Author:</u> Telephone: Australia UK: Twitter: @Jools_Physio. Twitter: @garethphysio1.

**Keywords:** Barriers, facilitators, menopause, physiotherapy, uncertainty, musculoskeletal

## Abstract

**Introduction:** Menopause typically begins in the 40s (perimenopause) with symptoms such as hot flushes, fatigue, and musculoskeletal pain affecting 80% of women. Declining hormones during menopause can reduce bone density, muscle strength, and joint health, contributing to musculoskeletal (MSK) pain, reported in up to 85% of women. Physiotherapists play a key role in the management of MSK pain, however, menopause-related symptoms such as fatigue and low self-efficacy can hinder physiotherapy engagement.

**Objectives:** To explore the experiences of menopausal women with musculoskeletal (MSK) pain attending physiotherapy and identify barriers and facilitators to engagement.

**Design:** A qualitative study utilising semi-structured interviews. Reflexive thematic analysis was performed.

**Setting:** A single NHS elective orthopaedic hospital in the UK.

**Participants:** 12 participants recruited via e-posters and staff support group

**Results:** Two main themes were identified: uncertainty regarding symptoms and considerations for physiotherapists. Common barriers to physiotherapy were fatigue, cognitive issues, low self-efficacy and negative language used by healthcare professionals. Participants emphasised being asked directly about their menopausal status and feeling listened to were facilitators for engagement. Individualised treatment strategies and education were reported as recommendations to foster rapport and engagement.

**Conclusions:** Menopausal women with MSK pain experience uncertainty about their symptoms and feel their needs are not fully met by physiotherapists. Direct enquiry, individualised care plans and education may improve engagement and enhance therapeutic relationships. Physiotherapists are well-positioned to support women through this important transition, but specific training may be required to enhance their knowledge and skills in treating menopausal women with MSK pain.

**Contribution of paper:**

- This is the first study to explore barriers and facilitators to physiotherapy for women with menopausal symptoms through their own perceptions and experiences
- Menopausal women with MSK pain often experience high levels of uncertainty regarding their intrusive symptoms
- Physiotherapists should consider direct questioning, education and individualised treatment plans when treating menopausal women with MSK pain
- Training programmes may be required to improve the knowledge and skills of physiotherapists treating menopausal women with MSK pain.

## 1. Introduction

Menopause represents the cessation of a woman’s periods and is a retrospective diagnosis after 12 months without menstruation. Historically linked to reproductive changes, it is now understood as a neuroendocrine process affecting brain function with an increased risk of Alzheimer’s disease.^1^ Menopause impacts 1.5 million women in the UK, 12% of the global population, and is projected to affect one billion women by 2025.^2^ Menopause typically starts in the 40s (perimenopause) with symptoms like hot flushes, fatigue, anxiety, ‘brain fog’ (cognitive impairment) and musculoskeletal pain, affecting 80% of women.^3^ These symptoms commonly affect health, social life, and work, with 1 in 10 UK women leaving jobs due to menopause.^4^

Declining hormone levels during menopause can reduce bone density, muscle strength, and joint health, contributing to MSK pain, which affects up to 70% of menopausal women.^5^ Women in this life stage are twice as likely as men to experience joint pain.^6^ As there is no surgical target for most persistent MSK pain experienced by menopausal women, physiotherapists play a crucial role in pain management.^7^ Clinical guidelines suggest that physiotherapists who treat people with persistent musculoskeletal pain should address physical and psychological aspects of their persistent pain.^8^

Many symptoms of menopause, including low self-efficacy, fatigue, and multi-joint pain, are known barriers to treatments delivered by physiotherapists.^9,10^ Therefore, menopausal women may be less likely to respond to interventions delivered by physiotherapists. There is a paucity of high-quality randomised controlled trials evaluating the effectiveness of interventions which aim to reduce the MSK pain experienced by menopausal women. Improved understanding of the barriers and facilitators to physiotherapist led treatments, from the perspective of menopausal women would provide valuable insights that have the potential to improve awareness of physiotherapists and shape future treatment interventions designed to treat menopausal women with persistent MSK pain.^11^ This study is the first to explore the barriers and facilitators to physiotherapy for women with menopausal symptoms through their own perceptions and experiences.

## 2. Methods

This study is reported in accordance with the consolidated criteria for reporting qualitative research (COREQ) checklist.^12^ See Appendix 1.

### 2.1 Qualitative approach and research paradigm

An interpretive hermeneutic phenomenological study was undertaken and is situated within a feminist world view.

### 2.2 Researcher Characteristics and Reflexivity

The principal researcher (PR) was a female in her forties, specialising in musculoskeletal physiotherapy with a special interest in menopause. The PR was known by some of the participants (n=10) due to her role as a physiotherapist within the Trust and her membership of the staff menopause support group from which participants were recruited.

### 2.3 Inclusion and exclusion criteria

Potential participants were included if they were assigned female gender at birth and still identified as female; they had accessed physiotherapy within the NHS for the treatment of MSK pain whilst experiencing menopausal symptoms; and were able to speak and understand the English language. People with conditions that exhibit symptoms like those of menopause were excluded, for example a history of inflammatory conditions, systemic/infective disease, or previous diagnosis of fibromyalgia or persistent multi-joint pain.^13^ People who were unable to be interviewed due to cognitive, speech or hearing problems were ineligible for participation. Whilst this study focused on cisgender women due to the existing research, it is acknowledged that menopausal symptoms are not exclusive to this population.

### 2.4 Context and setting

All interviews were undertaken in a private clinic room at a single NHS elective orthopaedic hospital in the UK, whether online or in person. Interviews were conducted solely between the PR and the participant, except for the first interview, which the second author attended to ensure consistency and minimise bias.

### 2.5 Sampling and sample size

A convenience sample of women were included, and sample size was determined by the concept of information power.^14^ Information power identified a lower number of participants needed due to the specificity of the sample, focus of the research question and context and the analysis strategy.

### 2.6 Procedure and data collection

Participants were recruited by the principal researcher (PR) via email to the Menopause staff support group and an e-poster circulated via the intranet within a single NHS Trust. Participants consented to a single, semi-structured interview online, via telephone or in person, dependent on participant preference. The interview schedule was developed by the research team, including a clinical academic who had experience of conducting interviews at the NHS trust based on the objectives of the study. The schedule was piloted with a member of the physiotherapy staff and no changes were made. The interview schedule consisted of ten questions covering 4 main topics: background to physiotherapy episode, experiences of menopause, attending physiotherapy whilst menopausal and reflections for the future (Appendix 2). Verbatim transcription was completed by the PR.

### 2.7 Data Analysis

Reflexive Thematic Analysis^15^ was undertaken. This method involved a six-phase analytical non-linear process: familiarisation with the data; generating codes; generating themes; reviewing themes; defining themes and writing up the data. Details of each stage of this step-by-step process are shown in Table 1. The second team member acted as a critical friend during the analysis process.

**Table 1:**
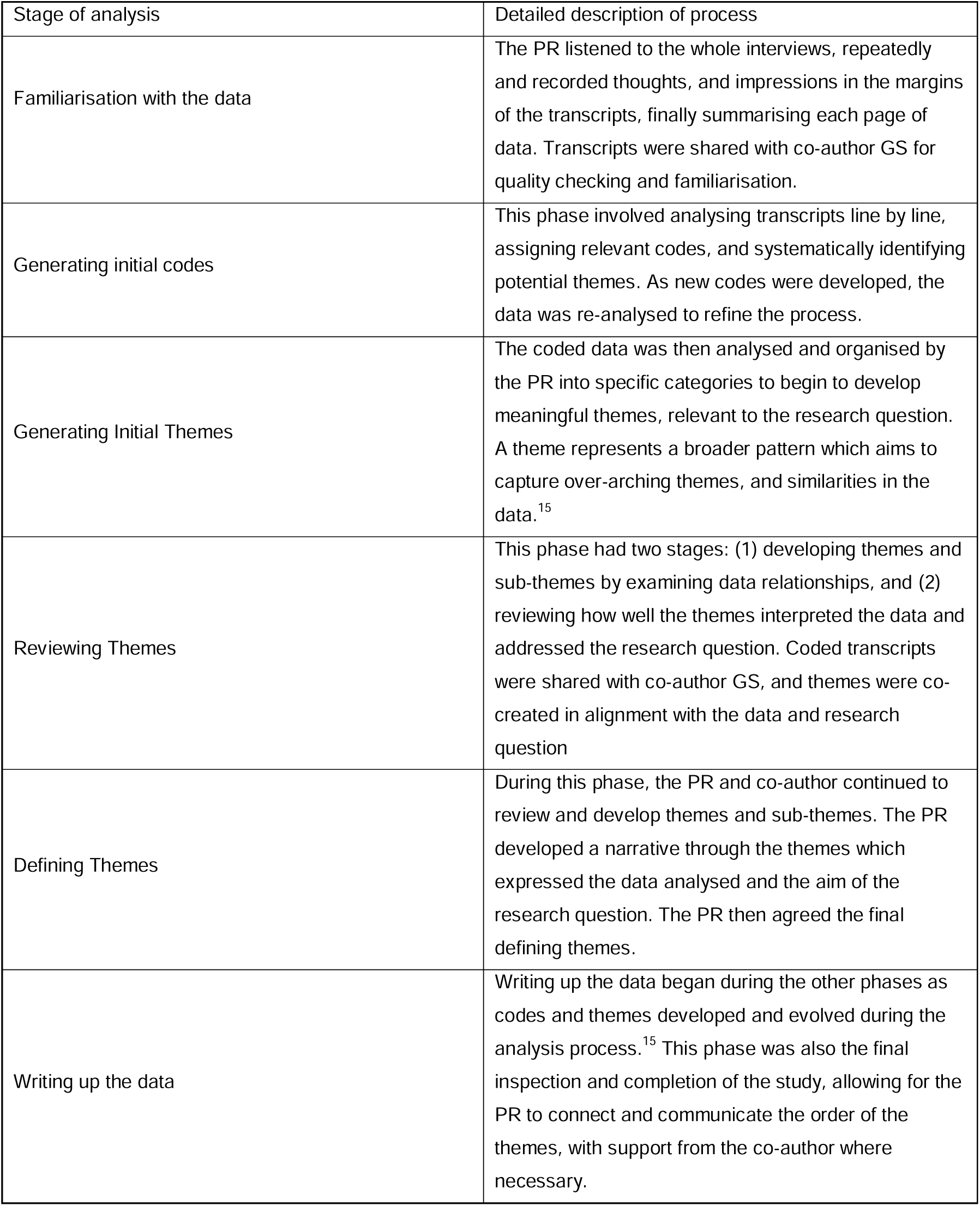
Stages of thematic analysis. ^15^

| Stage of analysis | Detailed description of process |
| --- | --- |
| Familiarisation with the data | The PR listened to the whole interviews, repeatedly and recorded thoughts, and impressions in the margins of the transcripts, finally summarising each page of data. Transcripts were shared with co-author GS for quality checking and familiarisation. |
| Generating initial codes | This phase involved analysing transcripts line by line, assigning relevant codes, and systematically identifying potential themes. As new codes were developed, the data was re-analysed to refine the process. |
| Generating Initial Themes | The coded data was then analysed and organised by the PR into specific categories to begin to develop meaningful themes, relevant to the research question. A theme represents a broader pattern which aims to capture over-arching themes, and similarities in the data.<sup>15</sup> |
| Reviewing Themes | This phase had two stages: (1) developing themes and sub-themes by examining data relationships, and (2) reviewing how well the themes interpreted the data and addressed the research question. Coded transcripts were shared with co-author GS, and themes were co-created in alignment with the data and research question |
| Defining Themes | During this phase, the PR and co-author continued to review and develop themes and sub-themes. The PR developed a narrative through the themes which expressed the data analysed and the aim of the research question. The PR then agreed the final defining themes. |
| Writing up the data | Writing up the data began during the other phases as codes and themes developed and evolved during the analysis process.<sup>15</sup> This phase was also the final inspection and completion of the study, allowing for the PR to connect and communicate the order of the themes, with support from the co-author where necessary. |

## 3. Results

### 3.1 Participants

A total of twelve participants (aged 49-59 years) agreed to participate in the research and none dropped out. Twelve semi-structured interviews were conducted by the PR either in-person (n=9), online via virtual platform Microsoft Teams (n=2) or recorded telephone call (n=1) and lasted between 23 and 60 minutes. All participants provided written consent and were also consented verbally prior to commencing the interview. At this point, they were advised that they may request to stop the interview at any time should they need to. No repeat interviews were carried out. Participant characteristics and presentation to physiotherapy are presented in Table 2.

**Table 2:** Characteristics of Participants.

| Participant Number | Age | Number of Physiotherapy Episodes | Duration of menopausal symptoms (years) | Presenting symptoms in physiotherapy (diagnosis provided where given) |
| --- | --- | --- | --- | --- |
| 1 | 50-55 | 3 | 7 | Knee pain, Gluteal tendinopathy, lower back pain |
| 2 | 50-55 | 2 | 1 | Ankle fracture/dislocation |
| 3 | 55-60 | 1 | 5 | Elbow pain, knee pain |
| 4 | 55-60 | 5 | 5 | Lower back pain, ankle fracture |
| 5 | 50-55 | 2 | 4 | Achilles tendinopathy |
| 6 | 50-55 | 3 | 5 | Lower back pain, frozen shoulder |
| 7 | 50-55 | 3 | 10 | Osteotomy-non-union |
| 8 | 50-55 | 2 | 2 | Knee pain-meniscal tear |
| 9 | 45-50 | 1 | 2 | Hip pain |
| 10 | 50-55 | 2 | 6 | Lower back pain, shoulder pain |
| 11 | 50-55 | 3 | 4 | Plantarfasciopathy, gluteal tendinopathy |
| 12 | 45-50 | Multiple over 15 years | 2 | Hip pain, total hip replacement |

### 3.2 Findings

A total of two over-arching themes were identified: Uncertainty regarding Symptoms and Considerations for Physiotherapists. Within these themes 4 sub-themes were identified; presentation to physiotherapy; experiences of menopause; barriers to physiotherapy; facilitators to physiotherapy. See table 3 for a summary of themes and example quotes.

**Table 3:** Summary of Themes and Example Quotes.

| Themes | Verbatim Quotes |
| --- | --- |
| 3.3.1: *Presentation to physiotherapy:*<br>Participants presentation to physiotherapy, including multiple episodes and multiple sites of pain | *'Is something else going off? So, it's just a vicious circle. You don't really know what's going on in your body (P6)'*<br>*'That's how I feel with menopause. Everything just hurts and you don't know which is gonna hurt today' P5*<br>*'It is hard to know if you are just getting old or if it's more, bits just start hurting' P1* |
| 3.3.2: *Experiences of menopause:*<br>Participants' beliefs and experiences of menopause | *'I hated this time of my life. I've not known who I am anymore. It's been difficult' (P10)*<br>*'I had a massive identity crisis. I couldn't recognise myself at all' (P2)*<br>*'I sit there in a puddle of tears thinking what is wrong with me' (P10)*<br>*'It made me panic that it was something serious, I must have cancer somewhere' (P12)*<br>*'It made me feel old and invisible for the first time in my life' (P2)*<br>*'Symptoms wise I have been a disaster. I'd never thought it could hit you as hard as it has hit me, I thought for a while, I'm just going mad' (P10)*<br>*'Words were gone. Things, people. People that I knew and have known for years, I couldn't remember their names' (P12)*<br>*'I knew it was a fridge, but I kept pointing at it saying I don't know what that is. The first thing you think is have I got dementia' P6*<br>*'I couldn't think. I couldn't sleep. I was trying to do simple tasks and* |
|  | *couldn't.' There was a couple of times where, I could have quite easily stepped in front of a car (P10)* |
| 3.4.1: *Barriers to Physiotherapy:*<br>Participants experiences of physiotherapy which led to lack of engagement and feeling dismissed and misunderstood | *'I think healthcare professionals are not very well trained in how to help so that has been very hit and miss' (P1)*<br>*'They just didn't listen' (P1)*<br>*'You're a female, over 50, what do you expect' (P2)*<br>*'They didn't ask me about menopause at all' (P7)* |
| 3.4.2: *Facilitators to Physiotherapy:*<br>Participants experiences of physiotherapy which reduced uncertainty, facilitated understanding of their symptoms | *' you feel vulnerable and different to how you have felt your whole life. It is about having conversations. Privately and with compassion' (P2)*<br>*'Like the foggy brain sometimes. He wrote it down for me so that when I got home, I could remember what I was supposed to be doing' (P5)*<br>*'it's about giving women access to knowledge and information about how to age well as a woman 'cause it's different for women' (P2)*<br>*'That is something with menopausal women that you need to consider. There is a lot going on with menopause' P7*<br>*'A recognition that it is individual as well. Different people will go through different things. It's not a one-size-fits-all' P3*<br>*'Asking that person what's easier for you? How would you manage this better?' P8* |

3.3. *Theme 1: Uncertainty Regarding Symptoms:*

This theme represented participants’ experiences and perceptions of their menopausal symptoms and how they presented to physiotherapy.

#### 3.3.1 sub-theme: Presentation to Physiotherapy

Participants attended physiotherapy with a variety of different MSK issues including gluteal tendinopathy, Achilles tendinopathy, shoulder pain and lower back pain. All but one participant had experienced multiple physiotherapy episodes. Several participants (n=6) self-reported multiple sites of MSK pain (however did not have a previous clinical diagnosis of persistent MSK pain). Whilst all participants attended physiotherapy for MSK related pain complaints, co-existing symptoms such as multiple sites of MSK pain and fatigue created uncertainty as to the diagnosis for their symptoms.

#### 3.3.2 *sub-theme:* Experiences of menopause

All participants highlighted that they were experiencing multiple symptoms associated with menopause at the point they consulted a physiotherapist, including multi joint pain, fatigue, brain fog, hot flushes, insomnia, weight gain and low mood. Symptoms are reported in table 4. Eleven participants reported that they felt that they had lost their confidence and identity, which negatively impacted their mental health and meaningful relationships. However, one participant did not report a change in self-concept. During initial presentation to physiotherapy and during treatment, all but one participant reported uncertainty regarding their symptoms in relation to whether they were due to menopausal changes, aging or something more sinister. Several participants reported that they were worried that they may be developing dementia, and one feared a cancer diagnosis. For some, this feeling of uncertainty was particularly distressing and isolating. Three participants were so distressed, they disclosed suicidal ideation.

**Table 4:** Symptoms of Menopause Reported by Participants.

| Menopausal Symptoms (as reported by participants) | Number of participants reporting symptom |
| --- | --- |
| Joint/muscle pain | 12 |
| Fatigue | 12 |
| Night sweats | 10 |
| Hot flushes | 9 |
| Brain fog | 10 |
| Anxiety and depression | 11 |
| Low mood | 11 |
| Sleep disturbance | 12 |
| Anger | 10 |
| Distress | 6 |
| Loss of confidence | 11 |
| Irritability | 11 |
| Mood swings | 12 |
| Forgetfulness | 10 |

Most participants reported a significant decrease in their quality of life, stating that their symptoms were intrusive and troubling. Some found that their symptoms made managing simple activities of daily living such as cooking and household chores extremely difficult. Many had to give up activities that brought them pleasure and improved their quality of life, secondary to their symptoms.

### 3.4 *Theme 2:* Considerations for Physiotherapists

This theme represents the insights shared by participants regarding the barriers and facilitators to physiotherapist led treatments.

#### 3.4.1 sub-theme: *Barriers to physiotherapy*

Eleven participants reported that they had had negative experiences with healthcare professionals in relation to menopause, including both GP’s and physiotherapists. Many participants felt that they had been dismissed based on their age and gender and had not been listened to effectively. Few participants were asked directly about their menopausal status and only one discussed menopause as a potential contributing factor to their symptoms.

Several participants reported that fatigue was a barrier to engaging in both the appointment with a physiotherapist and the home exercise programme they prescribed. ‘Brain fog’ (cognitive impairment) and variable MSK pain were also reported to be troublesome when attempting to engage in the home exercise component. Several participants stated that they felt they did not get individualised care, allowing for flexibility in exercises when they may be having a challenging day. Some participants reported that it may be difficult for some women to be asked about their menopause journey by a male physiotherapist, but most felt it was important to be asked regardless. Some participants felt that a female physiotherapist was more likely to ask direct questions about menopause than a male.

#### 3.4.2 sub-theme: *Facilitators of physiotherapy*

All participants were keen to discuss their menopause journey and welcomed direct questions from the physiotherapist. For most, privacy was a crucial component as some participants felt vulnerable when talking about their experiences. All participants reported that they felt heard and supported when asked direct questions. All participants reported the importance of being guided through their exercise programme in the session with the opportunity for flexibility in their plan. All felt a written or electronic plan available for home sessions was vital for engagement as this was helpful if they were experiencing cognitive issues such as brain fog or forgetfulness. Signposting to specialist menopausal services / support groups was also regarded as highly valuable for better knowledge and understanding.

## 4. Discussion

This study aimed to explore the experiences of menopausal women undergoing physiotherapy during the menopausal transition and identify barriers and facilitators to physiotherapy engagement. The findings underscore the significant, multidimensional impact of menopause on women’s quality of life. Participants commonly reported confusion and uncertainty about the nature and origin of their symptoms, such as musculoskeletal pain, fatigue, and low mood which presented as barriers to engagement in physiotherapy. Many struggled to distinguish menopausal changes from the natural aging process, which compounded feelings of distress and negatively impacted mental health and quality of life. Direct enquiry of menopausal status, listening with empathy and signposting to specialist services were all reported to be facilitators. Participants also stated that written or electronic exercise prescription was essential to mitigate against cognitive issues and having flexibility in their programme to allow for days which were more challenging.

Uncertainty in illness has historically been recognised as a critical psychological stressor.^16,17^ Although menopause is a natural physiological phenomenon rather than a pathological condition, its unpredictable and ambiguous symptoms often create fear, worry, and doubt among women.^18^ Studies have shown that higher levels of uncertainty are associated with more frequent and disturbing symptoms.^19^ This study highlights the uncertainty experienced by menopausal women who present to physiotherapists with MSK pain regarding both their pain and menopausal symptoms.

Several studies ^21,2,20^ highlight barriers for menopausal women such as motivation and accessibility to services. However, they do not consider the impact of uncertainty related to menopausal symptoms and musculoskeletal pain on engagement in physiotherapy intervention. Furthermore, the potential uncertainty experienced by the clinician may contribute to the lack of direct questioning, thus perpetuating further uncertainty for both clinician and the patient. Clinical uncertainty as a concept has been identified with increasing interest recently^22^ with further research highlighting how this may be addressed and supported moving forward.^23^ Recent evidence has demonstrated the value of educational intervention on uncertainty for menopause.^24^ Whilst diagnostic uncertainty pervades persistent MSK pain and menopausal symptoms, the acknowledgment of uncertainty and the accompanying complex emotions by clinicians has been shown to enhance patient-clinician communication, therapeutic relationships and provide reassurance to the patient.^25^ Given the above, further research evaluating the effectiveness of education programmes used to train physiotherapists to support menopausal women and educate clinicians could be highly valuable in MSK physiotherapy.

Participants highlighted numerous barriers to engaging in physiotherapy, including physical symptoms such as fatigue, musculoskeletal pain, and cognitive difficulties (e.g., brain fog). Additionally, stigma, embarrassment, and the perception that symptoms were a part of aging often led women to feel dismissed or misunderstood by healthcare professionals, including physiotherapists. These findings are consistent with a UK government survey^26^ revealing that 91% of women felt inadequately informed about menopause and 36% did not feel comfortable discussing their symptoms with healthcare providers. These findings are echoed by^2^ who found only 47% felt they had the tools to manage their symptoms. Such barriers not only limit access to effective treatment but also exacerbate the psychological and physical challenges faced by women during this important transition.^27,20^

Despite the barriers, all participants emphasised the importance of being asked direct yet empathetic questions about their menopausal status. They valued physiotherapists who demonstrated understanding and provided emotional support. Education and holistic care were identified as key facilitators, with participants expressing a desire for physiotherapists to address their symptoms in a comprehensive and individualised manner. Participants reported that individualised treatment plans were essential to holistic care and fostering trust in the therapeutic relationship, thus improving engagement.^28^ These findings are supported by recent research^27,20^ that identifies education and social support as important facilitators to health improvement.

Physiotherapists are well-positioned to empower women through education, guidance, and targeted interventions. For instance, previous evidence has identified the value of physiotherapy, exercise, and menopause-specific education to improve quality of life in a small cohort.^29^ Further research supports the value of yoga and pelvic floor muscle training on psychological and physical quality of life scores among women with menopausal symptoms.^30^

Education plays a crucial role in symptom management and healthcare engagement for menopausal women which highlights the influence of cultural and social factors on women’s experiences of menopause.^24^ Women’s experiences of menopause vary significantly based on intersecting factors such as socioeconomic status, race, ethnicity, education, employment, and access to healthcare. These factors do not exist in isolation but rather combine to shape women’s access to support, the severity of their symptoms, and their overall quality of life during this transitional phase.^27,31^ Women with lower levels of education are often less informed about menopause and available interventions leaving them more vulnerable to the uncertainty and psychological distress associated with menopause.^32,33^

This lack of knowledge perpetuates misinformation and contributes to health inequalities.^27^ Furthermore, studies indicate that limited awareness of treatment options leaves women feeling isolated and unsupported.^2^

Healthcare providers, including physiotherapists, can play a key role in bridging this gap by offering tailored education, guidance, and individualised care to women with menopausal symptoms experiencing MSK pain. Empowering women with knowledge not only supports them to manage their symptoms more effectively but also reduces feelings of uncertainty, enhances self-efficacy, and fosters a sense of control during this significant transition.^2,27^

### Implications

The following clinical implications should be considered:

1. Menopausal women with MSK pain often experience high levels of uncertainty regarding their intrusive symptoms
2. Physiotherapists should consider direct questioning, education, individualised treatment plans and privacy when treating menopausal women with MSK pain
3. Training programmes may be required to improve the knowledge and skills of physiotherapists treating menopausal women with MSK pain.

### Limitations

This study had several limitations. The principal researcher’s familiarity with some participants and personal experience of menopausal symptoms may have introduced bias. Reflexivity and investigator triangulation helped mitigate these risks. The small sample size can demonstrate inferential generalisability but not statistical generalisability. Future research should include larger, diverse populations to provide a broader understanding of menopause and its effects, supporting the development of more inclusive and effective care practices.

## 5. Conclusion

Menopause is not simply the cessation of a woman’s reproductive years but a significant, complex developmental transition and is therefore a significant public health issue, given its profound impact on women’s QoL. This study suggests that women feel uncertain around both their MSK pain and their menopausal symptoms and do not feel that their needs are met by physiotherapists. Women with menopausal symptoms suggest that direct enquiry, individualised treatment plans (which account for their menopausal symptoms such as brain fog and fatigue), education and privacy would support engagement in physiotherapy intervention. Evidence suggests that these interventions may empower women with MSK pain during the menopausal transition to foster a sense of control and develop strategies to navigate this transition positively, improving QoL. Whilst physiotherapists are uniquely positioned to play a key role in supporting women during this transition through holistic care, specific training programmes may be needed to develop their knowledge and skills in managing this population and creating a supportive, therapeutic environment

## Data Availability

All data produced in the present study are available upon reasonable request to the authors

## Acknowledgments

I would like to thank Harriet Robinson for her valuable contribution to the development of this idea and the Research and Development department and Charitable Fund at the Royal Orthopaedic Hospital NHS trust for supporting the study.

## Ethical Approval

Ethical approval for this study was granted by HRA and Health and Care Research Wales (HCRW) on April 17^th^, 2024, IRAS Project ID 340067. All participants gave written informed consent prior to data collection taking place.

## Appendices: Supplementary Data

### Appendix 1 Consolidated criteria for reporting qualitative studies (COREQ): 32-item checklist^12^

| No | Item | Guide questions/description | Page |
| --- | --- | --- | --- |
| **Domain 1:**<br>**Research team**<br>**and reflexivity** |  |  |  |
| Personal<br>Characteristics |  |  |  |
| 1. | Interviewer/facilitator | Which author/s conducted the interview or focus group? | 8 |
| j2. | Credentials | What were the researcher's credentials? *E.g. PhD, MD* | 5 |
| 3. | Occupation | What was their occupation at the time of the study? | 5 |
| 4. | Gender | Was the researcher male or female? | 5 |
| 5. | Experience and training | What experience or training did the researcher have? | 5 |
| Relationship with participants |  |  |  |
| 6. | Relationship established | Was a relationship established prior to study commencement? | 5 |
| 7. | Participant knowledge of the interviewer | What did the participants know about the researcher? *e.g. personal goals, reasons for doing the research* | 5 |
| 8. | Interviewer characteristics | What characteristics were reported about the interviewer/facilitator? *e.g. Bias, assumptions, reasons and interests in the research topic* | 5/15 |
| **Domain 2: study design** |  |  |  |
| Theoretical framework |  |  |  |
| 9. | Methodological orientation and Theory | What methodological orientation was stated to underpin the study? *e.g. grounded theory, discourse analysis, ethnography, phenomenology, content analysis* | 5 |
| Participant selection |  |  |  |
| 10. | Sampling | How were participants selected? *e.g. purposive, convenience, consecutive, snowball* | 6 |
| 11. | Method of approach | How were participants approached? *e.g. face-to-face, telephone, mail, email* | 7 |
| 12. | Sample size | How many participants were in the study? | 8 |
| 13. | Non-participation | How many people refused to participate or dropped out? Reasons? | 8 |
| Setting |  |  |  |
| 14. | Setting of data collection | Where was the data collected? *e.g. home, clinic, workplace* | 6 |
| 15. | Presence of non-participants | Was anyone else present besides the participants and researchers? | 6 |
| 16. | Description of sample | What are the important characteristics of the sample? *e.g. demographic data, date* | 6/8 |
| Data collection |  |  |  |
| 17. | Interview guide | Were questions, prompts, guides provided by the authors? Was it pilot tested? | 7 |
| 18. | Repeat interviews | Were repeat interviews carried out? If yes, how many? | 8 |
| 19. | Audio/visual recording | Did the research use audio or visual recording to collect the data? | 8 |
| 20. | Field notes | Were field notes made during and/or after the interview or focus group? | No |
| 21. | Duration | What was the duration of the interviews or focus group? | 8 |
| 22. | Data saturation | Was data saturation discussed? | 6 |
| 23. | Transcripts returned | Were transcripts returned to participants for comment and/or correction? | No |
| **Domain 3:**<br>**analysis and findings** |  |  |  |
| Data analysis |  |  |  |
| 24. | Number of data coders | How many data coders coded the data? | Table 1 |
| 25. | Description of the coding tree | Did authors provide a description of the coding tree? | Table 1 |
| 26. | Derivation of themes | Were themes identified in advance or derived from the data? | Table 1 |
| 27. | Software | What software, if applicable, was used to manage the data? | N/A |
| 28. | Participant checking | Did participants provide feedback on the findings? | No |

| Reporting |  |  |  |
| --- | --- | --- | --- |
| 29. | Quotations presented | Were participant quotations presented to illustrate the themes / findings? Was each quotation identified? *e.g. participant number* | Table 3 |
| 30. | Data and findings consistent | Was there consistency between the data presented and the findings? | 8 |
| 31. | Clarity of major themes | Were major themes clearly presented in the findings? | 8/9/10 |
| 32. | Clarity of minor themes | Is there a description of diverse cases or discussion of minor themes? | 8/9/10<br>Table 3 |

### Appendix 2: Interview Schedule

**<u>Interview Schedule Trial Number:</u>**

**<u>Screening Number:</u>**

**<u>Age:</u>**

**<u>Duration of physio intervention/no of appointments attended?</u>**

#### Background to physiotherapy episode

1. Tell me about the condition you went to physio with
2. What were the outcomes of your physiotherapy intervention?

#### Experiences of menopause

3. Can you share some of your experiences of menopause?
4. In what way does it affect your day-to-day life?

#### Attending physio whilst menopausal

5. Can you share some of your experiences of attending physiotherapy whilst experiencing menopausal symptoms?
6. What were your impressions of the treating physiotherapist? *Prompts:*
  ∘ *Do you have any examples of things which have influenced your impressions?*
  ∘ *Have your expectations been met by the physiotherapist?*
7. Do you feel that your physiotherapist addressed your menopausal symptoms as part of your treatment/discussion? *Prompts:*
  ∘ *Can you give examples of how?*
8. Can you share of your perception of the physiotherapists’ understanding of menopause?

#### Reflections for the future

